# Superior prognostic value of three-dimensional echocardiography-derived right ventricular ejection fraction: a meta-analysis

**DOI:** 10.1101/2022.06.02.22275907

**Authors:** Alex Ali Sayour, Márton Tokodi, Csilla Celeng, Richard A. P. Takx, Alexandra Fábián, Bálint K. Lakatos, Rocco Friebel, Elena Surkova, Béla Merkely, Attila Kovács

## Abstract

**Aims:** We aimed to confirm that three-dimensional echocardiography (3DE)-derived right ventricular (RV) ejection fraction (EF) is a more robust predictor of adverse cardiopulmonary outcomes than the conventional echocardiographic parameters.

**Methods and Results:** We performed a meta-analysis of studies reporting the impact of unit change of RVEF, tricuspid annular plane systolic excursion (TAPSE), fractional area change (FAC), and free-wall longitudinal strain (FWLS) on clinical outcomes (all-cause mortality and/or adverse cardiopulmonary outcomes). Hazard ratios (HR) were rescaled by the within-study standard deviations (SD) to represent standardized changes. Within each study, we calculated the ratio of HRs related to 1 SD reduction in RVEF versus TAPSE, or FAC, or FWLS, to quantify the predictive value of RVEF relative to the other metrics. These ratios of HRs were pooled using random-effects models.

Ten independent studies were identified as suitable, including data on 1,928 patients with various cardiopulmonary conditions. Overall, 1 SD reduction in RVEF was robustly associated with adverse outcomes (HR: 2.64 [95% CI: 2.18 to 3.20], p<0.001; heterogeneity: I^2^=65%, p=0.002). In studies reporting HRs for RVEF and TAPSE, FAC, or FWLS in the same cohort, RVEF had superior predictive value per SD reduction versus the other three parameters (vs. TAPSE, HR: 1.54 [95% CI: 1.04 to 2.28], p=0.031; vs. FAC, HR: 1.45 [95% CI: 1.15 to 1.81], p=0.001; vs. FWLS, HR: 1.44 [95% CI: 1.07 to 1.95], p=0.018).

**Conclusion:** 3DE-derived RVEF has superior prognostic value compared with conventional RV indices, therefore, it might further refine the risk stratification of patients with cardiopulmonary diseases.

**Graphical Abstract:** Added predictive value of three-dimensional (3D) echocardiography-derived right ventricular ejection fraction (RVEF) versus conventional metrics of RV systolic function on clinical outcomes: a meta-analysis of 10 studies. FAC: fractional area change, FWLS: free-wall longitudinal strain, HR: hazard ratio, SD: standard deviation, TAPSE: tricuspid annular plane systolic excursion

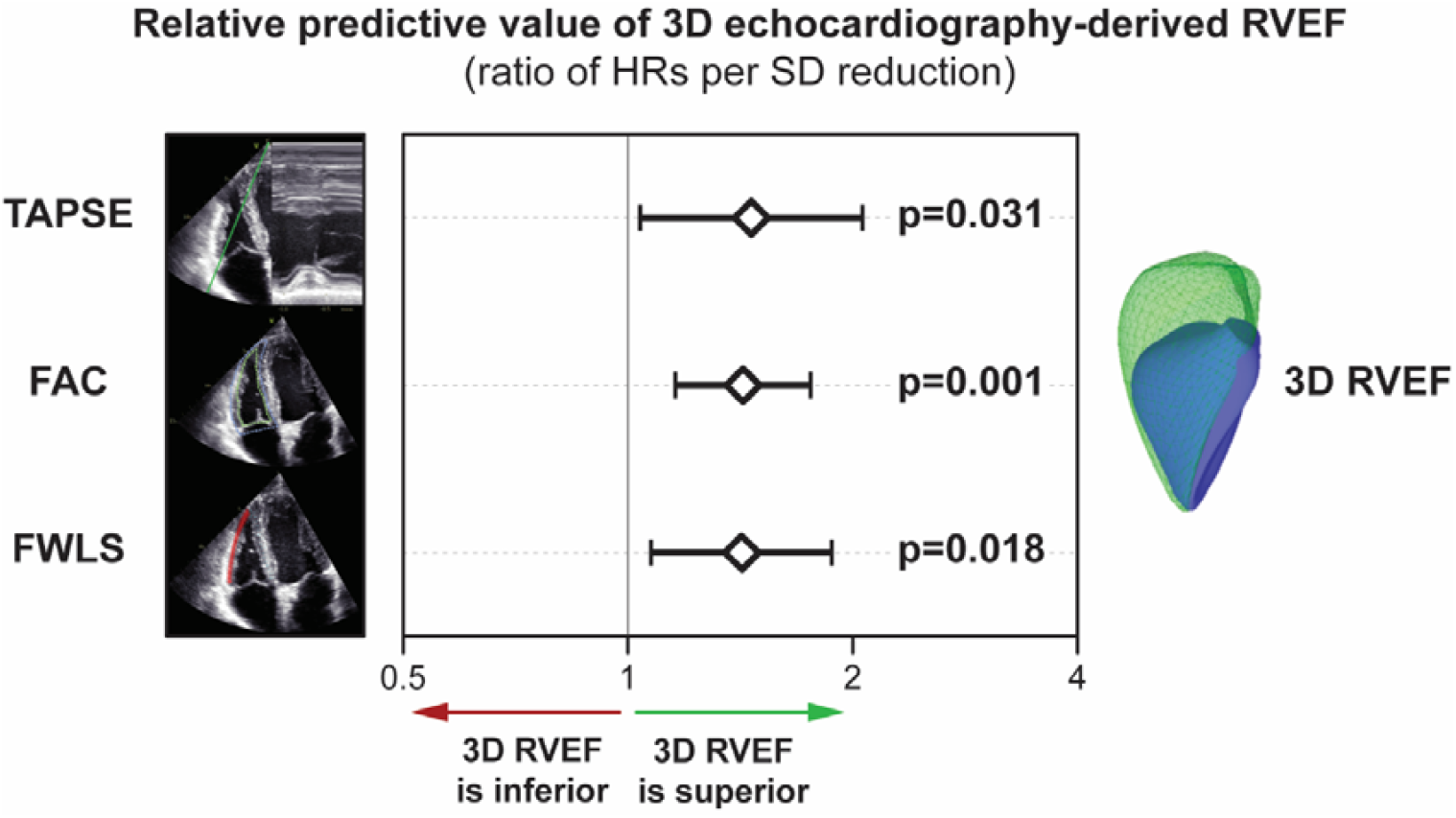

## INTRODUCTION

Assessment of right ventricular (RV) morphology and function is commonly included in comprehensive echocardiographic protocols of the clinical routine (1). RV dysfunction is closely associated with symptom burden and excess mortality in patients with various cardiopulmonary conditions, warranting efforts to discover the condition in the early stage of the disease (2).

The easily and routinely assessed parameters (e.g., tricuspid annular plane systolic excursion - TAPSE, fractional area change - FAC, free wall longitudinal strain - FWLS) can only partially portray the complex functional characteristics of the RV; therefore, they may fail to capture the full spectrum of RV dysfunction and associated adverse clinical outcomes (3).

Geometrically, the normal RV consists of a concave free wall surface and an opposing convex interventricular septum resulting in a crescent-shaped short-axis beyond the separated inflow and outflow parts. In that particular context, the M-mode and two-dimensional (2D) echocardiography-based measurements are rather simplistic approaches that do not account for such a complex three-dimensional (3D) shape, with the inherent risk of significant information loss. Moreover, from the functional aspect, recent data highlighted the importance of non-longitudinal mechanical components, which are entirely neglected or just partially reflected by conventional measures (4). 3D echocardiography-derived RV ejection fraction (RVEF) is a well-validated and reproducible parameter, which may overcome the shortcomings discussed above (5). Despite the physiological and technical advantages of RVEF measurement, it remains to be elucidated whether this translates into added prognostic value versus the conventional echocardiographic metrics.

Accordingly, we hypothesized that 3D echocardiography-derived RVEF is a more robust predictor of all-cause mortality and/or adverse cardiopulmonary outcomes than conventional echocardiographic parameters of RV systolic function.

## RESEARCH DESIGN AND METHODS

### Data Sources and Study Selection

This meta-analysis was conducted according to the Meta-analysis Of Observational Studies in Epidemiology (MOOSE) Guidelines (6). The study protocol was preregistered on PROSPERO (registration number: CRD42018110771). Two collaborators (M.T. and A.K.) independently assessed articles from PubMed and EMBASE from inception until 11 March 2022 using a predefined search strategy with the following inclusion criteria (Figure 1 and Supplementary Table 1): (1) English language studies published in peer-reviewed scientific journals; (2) studies reporting original investigations on human subjects; (3) adult age (>18 years) of all included participants; (4) studies with more than 20 subjects; (5) studies with 3D echocardiography performed and RVEF measured; (6) studies with all-cause mortality and/or adverse cardiopulmonary outcomes reported as hazard ratios (HRs, and 95% confidence intervals, CI) per unit change in 3D echocardiography-derived RVEF; and (7) at the same time, on the same cohort, reported at least one of the following RV functional parameters: TAPSE, FAC or FWLS. A manual reference check of eligible full-text articles was performed to identify studies missed by our systematic search. Disagreement was resolved by consensus. When separate publications from the same research group on seemingly overlapping cohorts were identified, the study involving the higher number of subjects was included in our final analysis.

**Figure 1.**
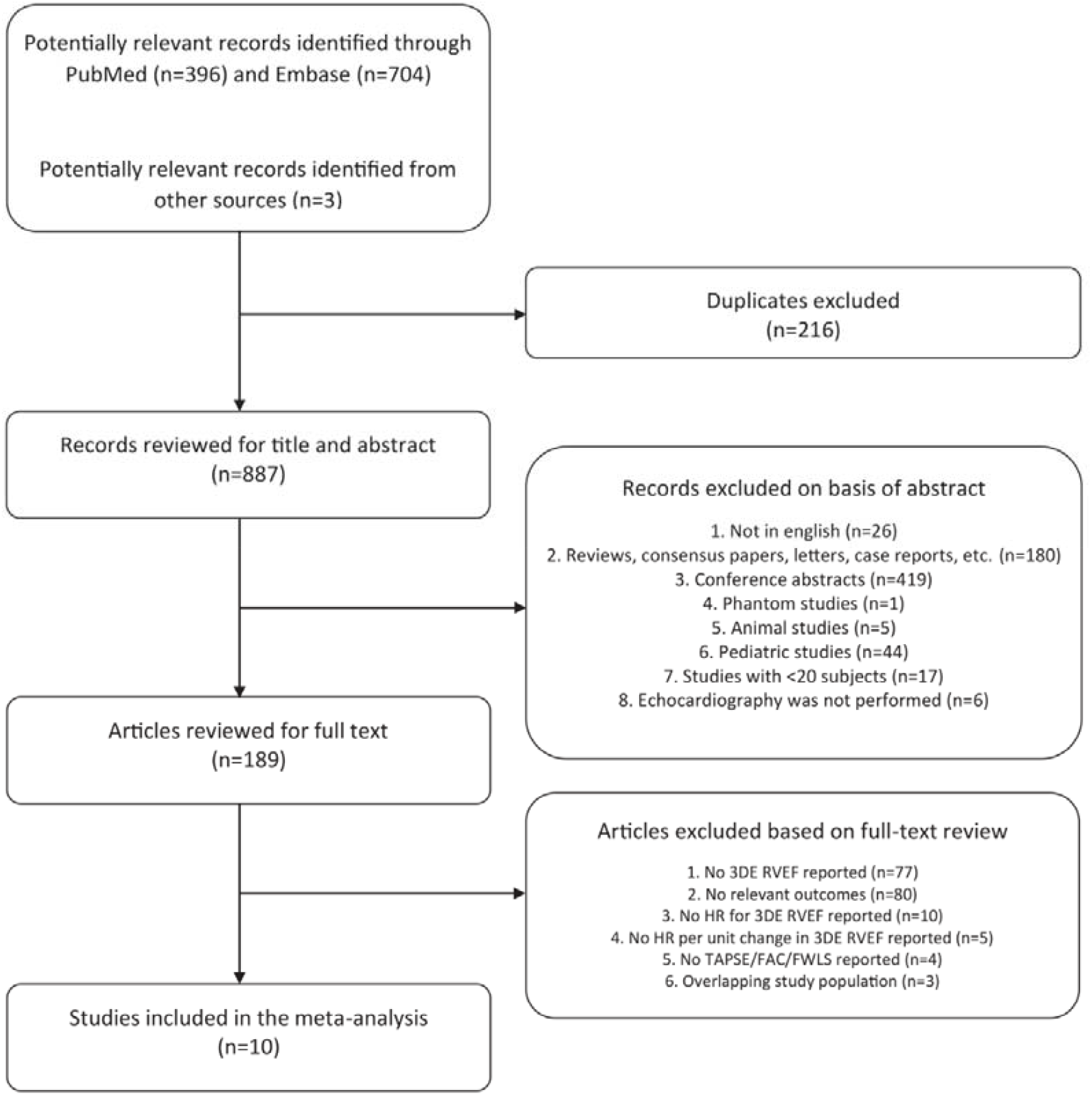
Study selection flowchart. 3DE: three-dimensional echocardiography, FAC: fractional area change, FWLS: free-wall longitudinal strain, HR: hazard ratio, RVEF: right ventricular ejection fraction, TAPSE: tricuspid annular plane systolic excursion

### Data Extraction and Quality Assessment

Data were extracted on study design, baseline characteristics of the cohorts, echocardiographic parameters, and the predefined outcomes for all included studies by two collaborators (M.T. and A.K.). Study quality was ascertained using the Quality In Prognosis Studies (QUIPS) tool in consensus (7).

### Data Synthesis and Analysis

HRs and respective 95% CIs reporting the association between the unit change of the prespecified echocardiography-derived RV functional parameters (RVEF and TAPSE, or FAC, or FWLS) and clinical outcomes were extracted from eligible publications. We limited our inclusion to studies that allocated HRs for RVEF and TAPSE, or FAC, or FWLS to the same endpoint within each study, as per the inclusion criteria. The majority of studies reported HRs and 95% CIs relative to 1 unit increase in 3D RVEF (1% increase), TAPSE (1mm increase), FAC (1% increase), and FWLS (1% increase in absolute value). Others reported these effect sizes per standard deviation (SD) change (8). To facilitate comparison of RVEF with TAPSE, FAC, and FWLS, all HRs and 95% CIs were rescaled by the within-study SD of the respective echocardiographic parameter to present a standardized change in the absolute value of each parameter (RVEF and TAPSE, or FAC, or FWLS) as previously described (9). Each SD reduction in the given echocardiographic parameter represents an increase in hazard, resulting in direct comparability of the predictive value of these parameters. Then, within each study, we calculated the ratio of HRs related to SD change in RVEF and TAPSE, FAC, or FWLS, to quantify the predictive value of RVEF relative to the other metrics. A ratio of >1.00 denotes that 1 SD reduction in RVEF is related to a greater hazard increment relative to 1 SD reduction in the other metric. The ratios of HRs related to each study were pooled using a random-effects model (DerSimonian-Laird). These pooled estimates represent the overall difference in predictive value of 1 SD reduction in RVEF versus 1 SD reduction in TAPSE, FAC, or FWLS, respectively. Forest plots were generated to visualize these differences. Statistical heterogeneity (referred to as heterogeneity) was assessed using the Cochran Q homogeneity test, and Higgins and Thompson I^2^ (10). The I^2^ heterogeneity was categorized as follows: 0-50% low, 50-75% moderate, >75% high. As a post hoc analysis using mixed-effects meta-regression, we explored whether follow-up duration, differences in baseline disease of cohorts (primary diagnosis of pulmonary hypertension vs. other cardiopulmonary conditions), or the type of endpoints (mortality only vs. composite) explained heterogeneity of the pooled estimates, yielding pseudo-R^2^ values (which refers to the percentage of heterogeneity explained by the given variable).

All statistical analyses were performed in Stata 17.0 (StataCorp LLC, College Station, TX, USA). A two-tailed p<0.05 was considered statistically significant.

### Sensitivity analyses

Funnel plots were constructed to visually inspect the small-study effect (corresponding to publication bias) according to each echocardiographic parameter and related clinical outcomes. The nonparametric Begg’s rank correlation test was used to quantify the association between the effect sizes and measures of precision (standard errors). Nonparametric trim-and-fill analysis as per Duval and Tweedie was performed to correct for the small-study effect using the R0 estimator (11). We used the DerSimonian Laird random-effects method for both the iteration and pooling steps during the trim-and-fill analyses.

## RESULTS

### Study selection and characteristics

A total of 189 articles were subject to full-text review. According to the predefined inclusion and exclusion criteria, 13 studies were found suitable. In three instances (including 3-3 studies), there was an apparent overlap between the patient cohorts (8, 12-16); therefore, studies with a higher number of participants were included (12, 14, 16). Overall, ten (12, 14, 16-23) independent studies were included in the final quantitative analysis (Figure 1), which reported the impact of unit change of RVEF and TAPSE [n=8] (12, 15, 17, 19-23), or FAC [n=7] (12, 18-23), or FWLS [n=7] (16, 18-23), on clinical outcomes (all-cause mortality and/or cardiopulmonary adverse events) as HRs. Four studies were prospective, while six were retrospective. Only three studies reported associations with all-cause mortality (12, 21, 23), the others reported composite cardiopulmonary endpoints (Table 1). We assessed the risk of bias within the studies using the QUIPS tool (Supplementary Figure 1).

**Table 1.**
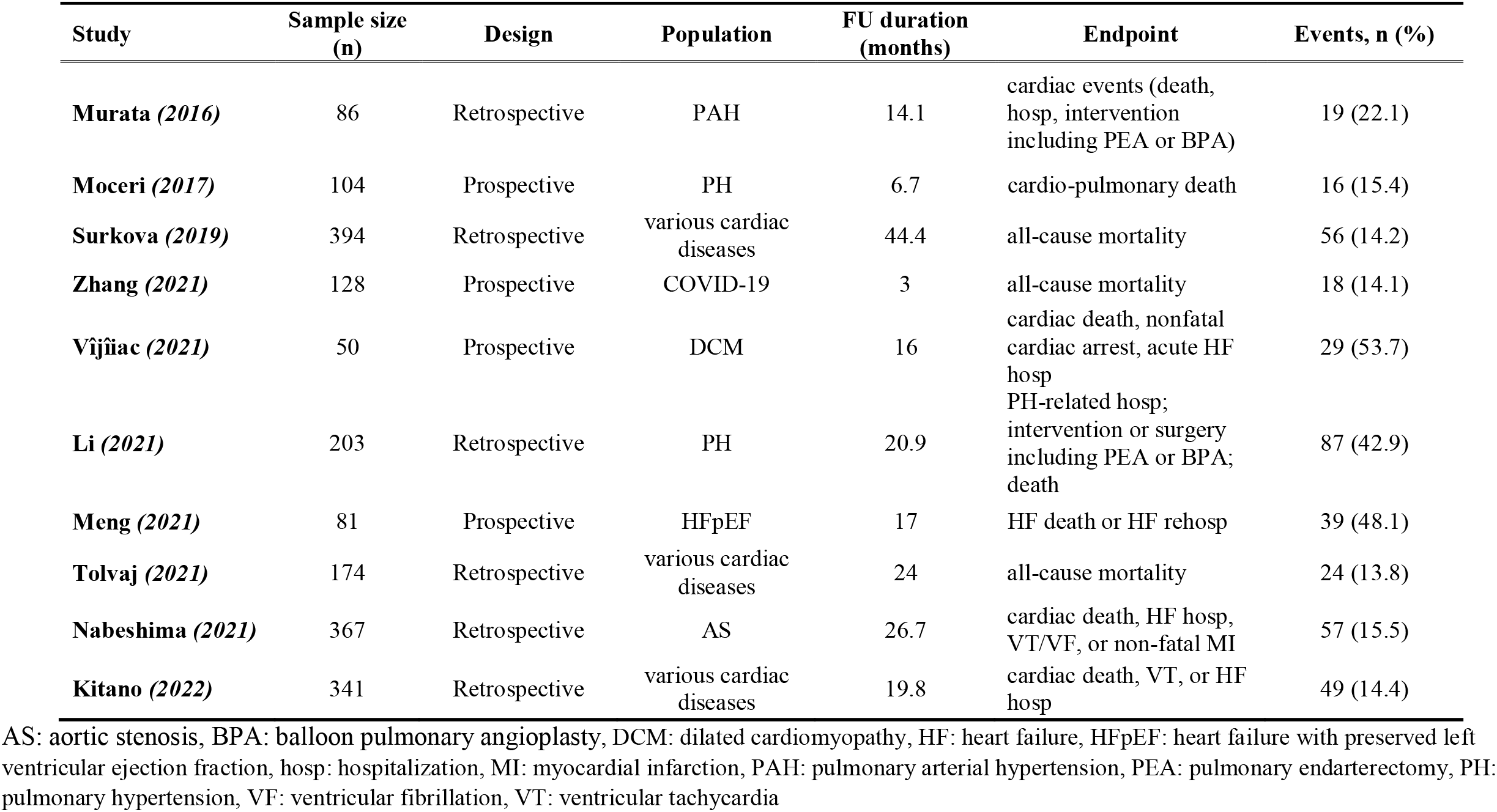
Study designs and clinical endpoints.

| Study | Sample size (n) | Design | Population | FU duration (months) | Endpoint | Events, n (%) |
| --- | --- | --- | --- | --- | --- | --- |
| **Murata (2016)** | 86 | Retrospective | PAH | 14.1 | cardiac events (death, hosp, intervention including PEA or BPA) | 19 (22.1) |
| **Moceri (2017)** | 104 | Prospective | PH | 6.7 | cardio-pulmonary death | 16 (15.4) |
| **Surkova (2019)** | 394 | Retrospective | various cardiac diseases | 44.4 | all-cause mortality | 56 (14.2) |
| **Zhang (2021)** | 128 | Prospective | COVID-19 | 3 | all-cause mortality | 18 (14.1) |
| **Vijñiac (2021)** | 50 | Prospective | DCM | 16 | cardiac death, nonfatal cardiac arrest, acute HF hosp | 29 (53.7) |
| **Li (2021)** | 203 | Retrospective | PH | 20.9 | PH-related hosp; intervention or surgery including PEA or BPA; death | 87 (42.9) |
| **Meng (2021)** | 81 | Prospective | HFpEF | 17 | HF death or HF rehosp | 39 (48.1) |
| **Tolvaj (2021)** | 174 | Retrospective | various cardiac diseases | 24 | all-cause mortality | 24 (13.8) |
| **Nabeshima (2021)** | 367 | Retrospective | AS | 26.7 | cardiac death, HF hosp, VT/VF, or non-fatal MI | 57 (15.5) |
| **Kitano (2022)** | 341 | Retrospective | various cardiac diseases | 19.8 | cardiac death, VT, or HF hosp | 49 (14.4) |

AS: aortic stenosis, BPA: balloon pulmonary angioplasty, DCM: dilated cardiomyopathy, HF: heart failure, HFpEF: heart failure with preserved left ventricular ejection fraction, hosp: hospitalization, MI: myocardial infarction, PAH: pulmonary arterial hypertension, PEA: pulmonary endarterectomy, PH: pulmonary hypertension, VF: ventricular fibrillation, VT: ventricular tachycardia

### Echocardiographic measurements

All study subjects underwent standard echocardiographic examination by experienced sonographers using commercially available ultrasound scanners (Supplementary Table 2). RVEF was measured by a single commercially available software (TomTec 4D RV-Function, TomTec Imaging GmbH, Unterschleissheim, Germany; reported version 2.0 or newer, standalone or embedded into another vendor’s platform). In the vast majority of the cases, an RV-focused apical window was used to acquire the full-volume 3D echocardiographic dataset using multi-beat reconstruction, which was finally utilized to measure RVEF. The initial semi-automated 3D contouring was further corrected manually by the investigators. TAPSE was measured using an M-mode recording, while FAC was assessed by contouring the end-diastolic and end-systolic RV endocardial surfaces in accordance with current guidelines (1). Two-dimensional free wall longitudinal strain was measured using commercially available software packages from two vendors (Supplementary Tables 2 and 3).

### Patient characteristics

The ten studies comprised data on 1928 patients. Clinical characteristics and definitions of composite endpoints are shown in Tables 1 and 2. The mean (±SD) age of the patient population was 63±15 years, 46% were females, and the follow-up duration ranged from 3 to 44 months. Three studies included patients with pulmonary hypertension exclusively (14, 16, 20), one included patients with COVID-19 only (23), one included patients with dilated cardiomyopathy only (22), one included patients with aortic stenosis only (18), one included heart failure patients with preserved left ventricular (LV) EF (HFpEF) only (19), and the remaining three studies included populations with a mixture of cardiovascular diseases (12, 17, 21).

**Table 2.** Demographic and clinical characteristics of the study populations.

| Study | Age, years | Female,<br>n (%) | BMI, kg/m<sup>2</sup> | BSA, m<sup>2</sup> | HR, bpm | SBP, mmHg | DBP,<br>mmHg | DM,<br>n (%) | HTN,<br>n (%) | DLP,<br>n (%) | CAD,<br>n (%) | CKD,<br>n (%) |
| --- | --- | --- | --- | --- | --- | --- | --- | --- | --- | --- | --- | --- |
| **Murata (2016)** | 50±17 | 63 (72) | NA | 1.6±0.2 | 72±14 | 111±17 | 64±13 | NA | NA | NA | NA | NA |
| **Moceri (2017)** | 66 (62-69) | 58 (56) | NA | 1.72±0.2 | NA | NA | NA | NA | NA | NA | NA | NA |
| **Surkova (2019)** | 57 (42-69) | 136 (35) | 24.5 (22.1-26.8) | 1.81 (1.67-1.95) | 68 (59-77) | 120 (110-130) | 70 (70-80) | 51 (12.9) | 190 (48.2) | 143 (36.3) | 119 (30.2) | NA |
| **Zhang (2021)** | 61±13 | 80 (50) | NA | 1.67±0.15 | 86 (80-99) | 130 (120-140) | 80 (73-88) | 18 (14.1) | 52 (40.6) | NA | NA | 1 (0.8) |
| **Vijñiac (2021)** | 61±14 | 16 (32) | NA | NA | 79±16 | 124±13 | 75±11 | 9 (18) | 34 (68) | NA | 0 (0) | NA |
| **Li (2021)** | 49±15 | 146 (72) | NA | 1.7±0.2 | NA | NA | NA | NA | NA | NA | NA | NA |
| **Meng (2021)** | 62±12 | 18 (35) | 25.5±3.6 | NA | 72.4±11.5 | 136.3±22.6 | 83.0±14.9 | 36 (44.4) | 46 (56.8) | NA | NA | NA |
| **Tolvaj (2021)** | 62±14 | 48 (28) | NA | 1.9±0.2 | NA | 126.2±19.7 | 74.7±16.5 | 39 (22.5) | 113 (65.3) | NA | 38 (22) | NA |
| **Nabeshima (2021)** | 77±10 | 199 (54) | 22.7±3.9 | 1.52±0.21 | 69.5±12.6 | 147.5±23.5 | 75.0±13.5 | 115 (31) | 295 (80) | NA | 79 (22) | 174 (47) |
| **Kitano (2022)** | 68 (58-76) | 115 (34) | NA | 1.62 (1.50-1.75) | 67 (59-76) | 127 (112-145) | 71 (63-79) | 101 (30) | 191 (56) | 149 (44) | 143 (42) | 149 (44) |

Data are presented as mean±SD or median (interquartile range). BMI: body mass index, BSA: body surface area, CAD: coronary artery disease, CKD: chronic kidney disease, DBP: diastolic blood pressure, DLP: dyslipidemia, DM: diabetes mellitus, HR: heart rate, HTN: systemic arterial hypertension, SBP: systolic blood pressure

### Outcomes

Table 1 contains the definitions of endpoints assessed in each included study. Among the 1928 patients, 394 (20.4%) reached the endpoint of all-cause mortality and/or adverse cardiopulmonary events.

In the ten studies, 1 SD reduction in RVEF was associated with a 2.64-fold (95% CI: 2.18 to 3.20, p<0.001) increase in the risk of all-cause mortality and/or adverse cardiopulmonary events (Figure 2). The moderate heterogeneity (I^2^=65%, p=0.002) found across studies was not explained by differences between follow-up duration (pseudo-R^2^=2%, p=0.062), endpoint definitions (pseudo-R^2^=0%, p=0.806), or primary diagnosis of pulmonary hypertension (pseudo-R^2^=0%, p=0.524). The funnel plot (Supplementary Figure 2) and Begg’s test for small-study effects (z=-1.43, p=0.15) showed that risk of publication bias was low. Accordingly, the trim-and-fill analysis (Supplementary Figure 3) showed that even if significant one-tailed publication bias occurred that favored the publication of highly positive studies (suggesting that RVEF is a powerful predictor of clinical outcomes), our pooled study estimate would not have been significantly altered (adjusted HR: 2.32 [95% CI: 1.86 to 2.90]).

**Figure 2.**
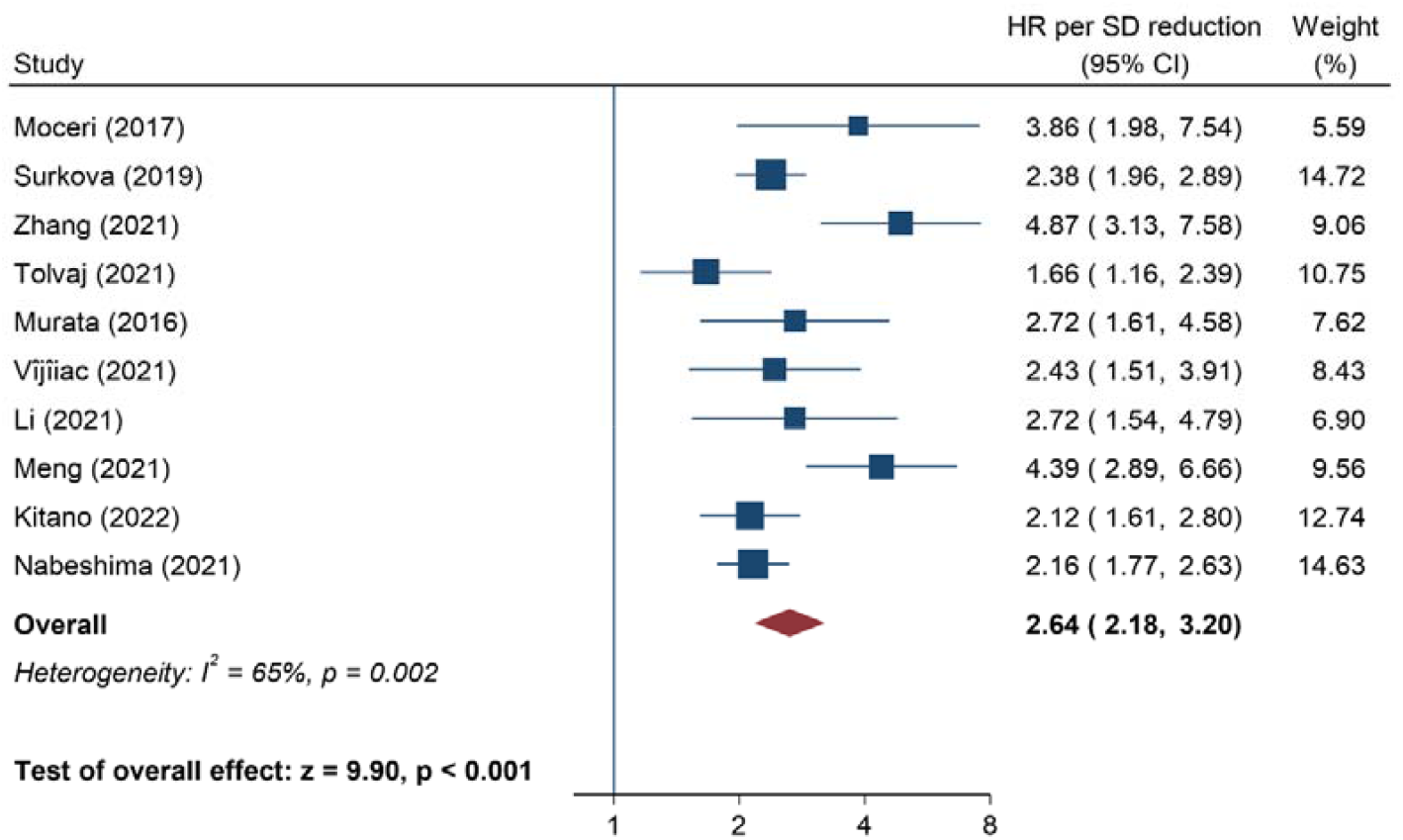
Three-dimensional echocardiography-derived right ventricular ejection fraction (RVEF) as predictor of all-cause mortality and/or composite adverse cardiopulmonary endpoints. The hazard ratios (HR) are per 1 standard deviation (SD) reduction in RVEF according to each study. Accordingly, a HR of >1.00 means that 1 SD reduction in RVEF is associated with increased risk of adverse events. CI: confidence interval

In studies reporting HRs for RVEF and TAPSE simultaneously in the same cohort, the SD reduction in RVEF (HR: 2.76 [95% CI: 2.16 to 3.54]) and TAPSE (HR: 1.81 [95% CI: 1.43 to 2.28]) were both significantly associated with adverse clinical outcomes (Supplementary Figure 4). However, the HR per SD change for RVEF as a correlate of outcomes was 1.54 (95% CI: 1.04 to 2.28, p=0.031) times greater than that of TAPSE, with moderate heterogeneity (I^2^=74%, p<0.001) (Figure 3). The latter was not related to study differences in follow-up duration (pseudo-R^2^=17%, p=0.15), endpoint definitions (pseudo-R^2^=0%, p=0.95), or primary diagnosis of pulmonary hypertension (pseudo-R^2^=0%, p=0.68). Begg’s test for small-study effects (z=0.62, p=0.54) indicated no evidence of substantial one-sided publication bias (Supplementary Figure 5).

**Figure 3.**
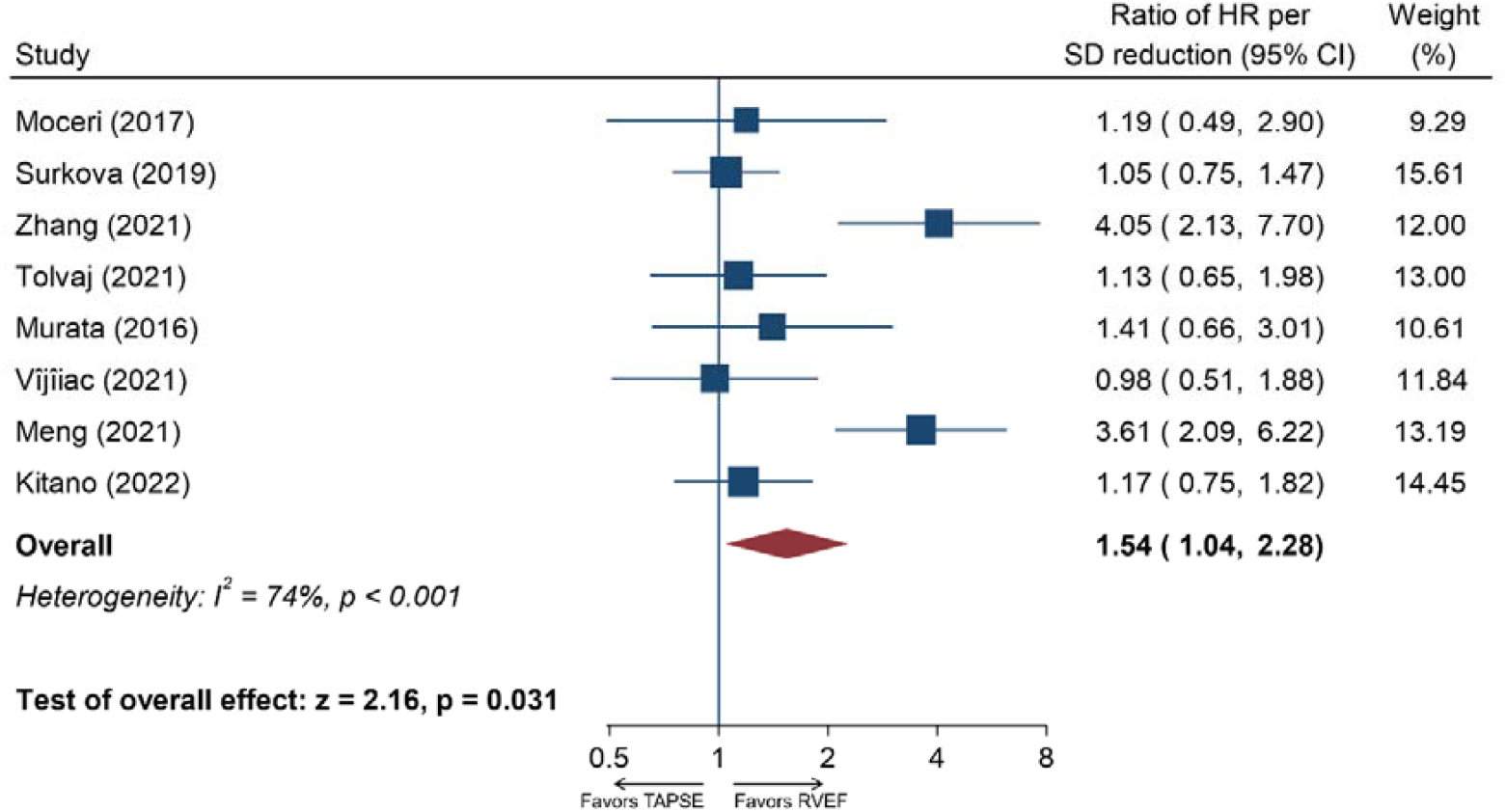
Three-dimensional echocardiography-derived right ventricular ejection fraction (RVEF) versus tricuspid annular plane systolic excursion (TAPSE) as predictors of all-cause mortality and/or composite adverse cardiopulmonary endpoints. The ratios of hazard ratios (HR) per 1 standard deviation (SD) reduction in RVEF/TAPSE are depicted. Accordingly, a ratio of HR >1.00 means that 1 SD reduction in RVEF is associated with a higher risk of adverse events compared to 1 SD reduction in TAPSE. CI: confidence interval

We found that 1 SD reduction in RVEF (HR: 2.68 [95% CI: 2.09 to 3.42]) and FAC (HR: 1.71 [95% CI: 1.44 to 2.02]), respectively, were associated with adverse outcomes in patients with various diseases (Supplementary Figure 6). The above HR per SD reduction in RVEF translates into 1.45 (95% CI: 1.15 to 1.81, p=0.001) times greater risk of adverse outcomes as compared to that of FAC (Figure 4). In this analysis, heterogeneity was low (I^2^=39%, p=0.13), to which differences in follow-up duration (pseudo-R^2^=0%, p=0.39), endpoint definitions (pseudo-R^2^=0%, p=0.22), and primary diagnosis of pulmonary hypertension (pseudo-R^2^=0%, p=0.89) had no significant contribution. The presence of publication bias in this analysis was not supported by the visual inspection of the funnel plot (Supplementary Figure 7) and Begg’s test (z=0.30, p=0.76).

**Figure 4.**
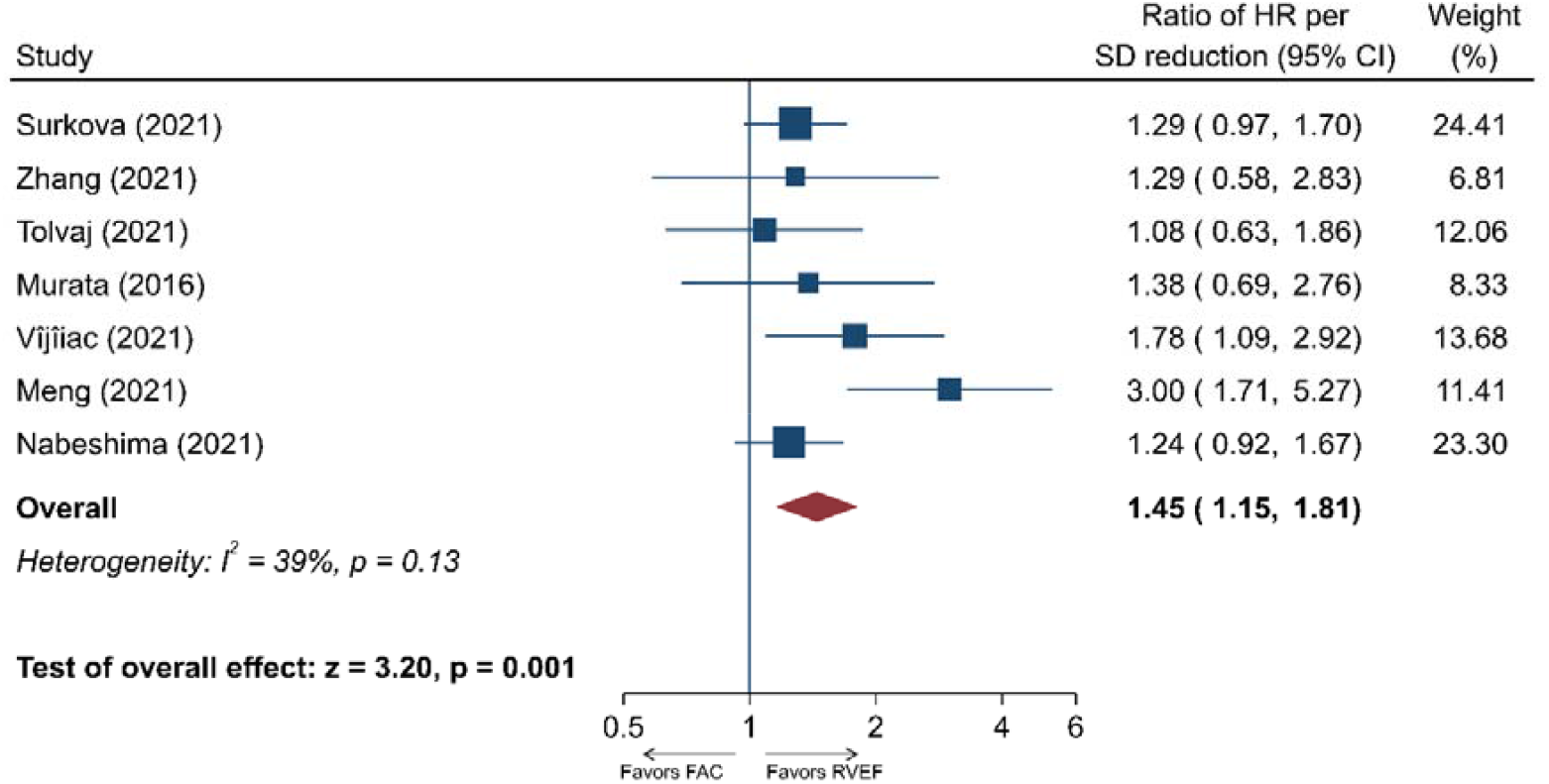
Three-dimensional echocardiography-derived right ventricular ejection fraction (3D RVEF) versus fractional area change (FAC) as predictors of all-cause mortality and/or composite adverse cardiopulmonary endpoints. The ratios of hazard ratios (HR) per 1 standard deviation (SD) reduction in RVEF/FAC are shown. Accordingly, a ratio of HR >1.00 means that 1 SD reduction in RVEF is associated with a higher risk of adverse events compared to 1 SD reduction in FAC. CI: confidence interval

Finally, in studies reporting the association of unit change in RVEF and FWLS on clinical outcomes in the same cohort, we found that 1 SD decrease in these parameters was significantly associated with adverse events (RVEF, HR: 2.76 [95% CI: 2.06 to 3.70]; FWLS, HR: 1.77 [95% CI: 1.42 to 2.21]) (Supplementary Figure 8). However, the strength of effect for the HR per SD reduction for RVEF was 1.44 (95% CI: 1.07 to 1.95, p=0.018) times higher than that of FWLS, suggesting a superior predictive value (Figure 5). None of the investigated factors (follow-up duration: pseudo-R^2^=0%, p=0.96; endpoint differences: pseudo-R^2^=0%, p=0.18; primary diagnosis of pulmonary hypertension: pseudo-R^2^=0%, p=0.60) contributed significantly to the low level of heterogeneity (I^2^=47%, p=0.08). No substantial small study effect was present based on the funnel plot (Supplementary Figure 9) and Begg’s test (z=0.30, p=0.76).

**Figure 5.**
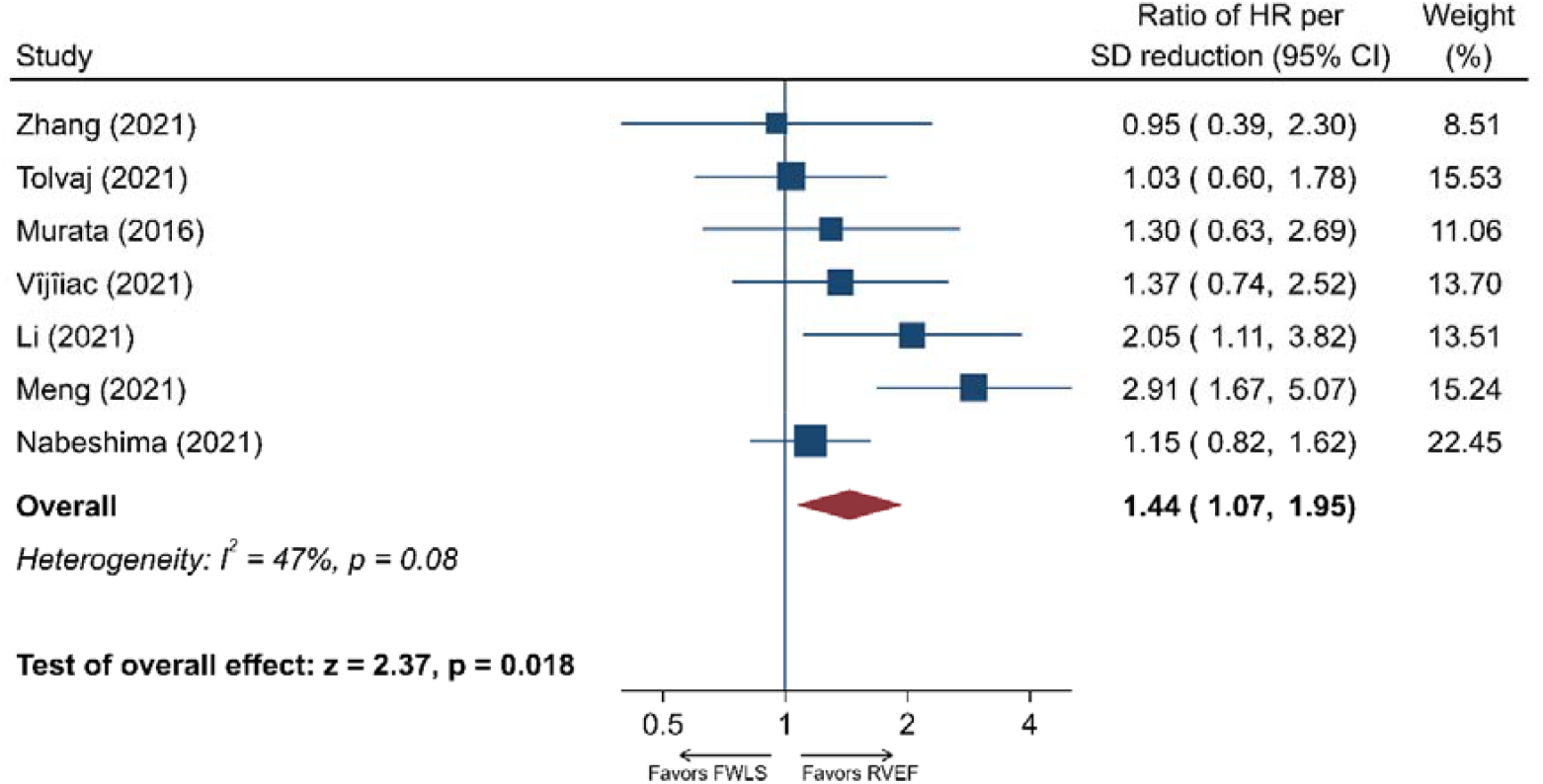
Three-dimensional echocardiography-derived right ventricular ejection fraction (3D RVEF) versus free-wall longitudinal strain (FWLS) as predictors of all-cause mortality and/or composite adverse cardiopulmonary endpoints. The ratios of hazard ratios (HR) per 1 standard deviation (SD) reduction in RVEF/FWLS are depicted. Accordingly, a ratio of HR >1.00 means that 1 SD reduction in RVEF is associated with a higher risk of adverse events compared to 1 SD reduction in FWLS. CI: confidence interval.

## DISCUSSION

In this meta-analysis, we found that RV dysfunction is a robust predictor of all-cause mortality and adverse cardiopulmonary outcomes in patients with various cardiopulmonary diseases. All four investigated echocardiographic parameters of RV systolic function were associated with these endpoints; however, 1 SD reduction in 3D echocardiography-derived RVEF was a significantly stronger predictor of adverse events compared with the same change in TAPSE, FAC, or FWLS (Graphical Abstract).

### Clinical significance of RV systolic dysfunction

Studies over the last decade have identified RV dysfunction as a correlate of symptom burden and a powerful predictor of adverse outcomes not only in right-sided heart diseases but also in conditions affecting the left ventricle primarily (24). In fact, the prevalence of RV dysfunction in patients with heart failure (HF) with reduced left ventricular EF (HFrEF) is 47%, being independently associated with excess mortality and HF admissions (25). Furthermore, one-third of patients with HFpEF present with a significant RV dysfunction (26). The presence of RV systolic dysfunction alongside the diagnosis of HFpEF harbors a ∼6-times higher risk of two-year mortality compared with the absence of it (26). Therefore, timely identification of RV dysfunction using sensitive and reliable functional parameters might enable a more sophisticated risk stratification in the clinical setting.

### Role of echocardiography in the assessment of RV function

According to two recent surveys, the most commonly used echocardiographic parameters to assess RV systolic function in the clinical routine is the M-mode TAPSE, followed by tissue Doppler imaging-derived S’ and FAC (27, 28). There is evidence that RV dysfunction is diagnosed in 37% of HFpEF patients by TAPSE, whereas FAC identifies a considerably lower fraction of cases [26%] (29). A meta-analysis also reported vast differences in the identification of RV dysfunction across parameters, with TAPSE suggesting RV dysfunction in 31% of HFpEF subjects, compared with 26% and 13% by S’ and FAC, respectively (30). These considerable diagnostic dissimilarities between conventional echocardiography-derived indices of RV function might stem from the complex 3D anatomy and distinct contraction patterns of the RV (13, 14). Therefore, a unifying RV functional parameter that can circumvent such limitations would be ideal for quantifying RV dysfunction and associated clinical risk in various pathological conditions. While 2D speckle-tracking echocardiography might overcome many limitations of conventional parameters by being less dependent on the angle of insonation and RV loading conditions (31), it still represents a single 2D tomographic plane. On the contrary, 3D echocardiography maps the entire endocardial surface of the RV independent of any assumption about its shape and function, providing an integrative parameter of RV systolic function – RVEF (24). Nonetheless, it is currently unclear whether 3D echocardiography-derived RVEF outperforms other RV function metrics in predicting adverse clinical outcomes.

### Prognostic significance of different echocardiographic parameters of RV systolic function

The present meta-analysis demonstrated that 3D echocardiography-derived RVEF was a robust predictor of adverse outcomes. We estimated that 1 SD reduction in RVEF conferred a ∼2.6 times higher risk of all-cause mortality and/or adverse cardiopulmonary events in a broad spectrum of patients with various cardiopulmonary conditions. This predictive value was unaffected by whether the population consisted of patients with a primary diagnosis of pulmonary hypertension. Therefore, our meta-analysis extends previous studies by showing that RV dysfunction forecasts adverse clinical events not only in patients with pulmonary hypertension (32) but also in those with HF, COVID-19, and aortic stenosis. Furthermore, our estimate was not affected by differences in follow-up durations, whether the endpoint included all-cause mortality only or other composite cardiopulmonary endpoints. Finally, we estimated that even if substantial publication bias occurred, our estimate would not significantly change.

As for the other parameters, we calculated that TAPSE, FAC, and FWLS are also predictors of adverse clinical outcomes, respectively. In fact, 1 SD reduction in each of these parameters was associated with a 1.71-1.81 times higher risk of unfavorable events. Therefore, RV dysfunction is a solid predictor of clinical outcomes irrespective of the echocardiographic parameter used. However, in a head-to-head comparison, we found that 1 SD reduction in RVEF conferred a 1.44-1.54 times higher risk of adverse outcomes as compared with a similar reduction in TAPSE, FAC, and FWLS, respectively, in the very same patient population. Consequently, 3D echocardiography-derived RVEF might identify a broader spectrum of patients at risk with RV dysfunction, in contrast with the other parameters, rendering it a more valuable tool to stratify patients with various cardiopulmonary conditions. This might translate into a timely identification of high-risk patients, paving the way for early effective countermeasures.

In the head-to-head comparison, RVEF remained a significantly better predictor of adverse clinical outcomes irrespective of whether the studied population included patients with pulmonary hypertension only or not. Therefore, RVEF might also overcome the limitations of TAPSE, FAC, and FWLS in primary left-heart diseases. Furthermore, differences in endpoint definitions and follow-up durations had no significant impact on the superiority of RVEF as a predictor of adverse outcomes. In a similar attempt to assess the pooled prognostic significance of different parameters of LV systolic function, a previous meta-analysis showed that LV global longitudinal strain (GLS) had a superior predictive value compared to LVEF (9). Notably, in our present meta-analysis focusing on RV function, RVEF was shown to outperform the predictive power of FWLS. This important finding may be attributable to the fact that RV FWLS reflects only one mechanical component (longitudinal shortening) in a single 2D tomographic plane (unlike three planes for LV GLS assessment) and, therefore, may not be an adequate representation of the contraction pattern of the complex 3D structure of the RV, especially under different pathophysiological conditions.

Overall, the results of our current meta-analysis support the broader implementation of 3D echocardiography for the assessment of RV systolic function in patients with cardiopulmonary disorders – irrespective of the primary site of the disease. Importantly, the use of contemporary automated 3D software solutions may even result in shorter analysis times compared with routine 2D evaluations (33). As the prognostically most powerful index of RV function, the inclusion of RVEF in everyday clinical decision-making and risk stratification models seems justified by the current knowledge base. Moreover, the assessment of RVEF as a trigger for specific therapies may also lead to clinical benefits for patients in the future.

## LIMITATIONS

The validity of the present meta-analysis is subject to the quality of the reporting of the included studies, rendering our findings hypothesis-generating only. First, the studies included in this meta-analysis were non-uniform in design, varied in the inclusion criteria, patients’ populations, echocardiographic equipment, technical aspects of 3D RV data acquisition, duration of follow-up, and definition of endpoints. Therefore, we opted for using a random-effects meta-analysis and performed a mixed-effects meta-regression to estimate the contribution of select factors (differences in patient populations, follow-up, and endpoint definitions) to the observed results. Second, not all 10 studies included in the current meta-analysis provided quantification of 3D echocardiography-derived RVEF and all three other RV parameters of interest at the same time, which led to a smaller number of studies included in each comparison (8 for RVEF vs. TAPSE, 7 for RVEF vs. FAC, and 7 for RVEF vs. FWLS). Third, as prespecified, due to a considerably lower amount of available and comparable data, tissue Doppler-derived tricuspid annular S’ velocity and RV GLS were not included in our analysis. While the former measures longitudinal shortening exclusively, the predictive value of the latter has recently been shown to be inferior compared to FWLS in HFrEF (34).

## CONCLUSIONS

Reduction in RV systolic function is robustly associated with adverse clinical outcomes in patients with various cardiopulmonary conditions. 3D echocardiography-derived RVEF identifies a broader spectrum of patients at risk than other RV systolic functional parameters. Therefore, RVEF might further refine the risk stratification of patients and inform clinical decision-making, potentially facilitating timely interventions.

## Data Availability

All data produced in the present study are available upon reasonable request to the authors.

## ABBREVIATIONS

2D: two-dimensional
3D: three-dimensional
CI: confidence interval
FAC: fractional area change
FWLS: free wall longitudinal strain
GLS: global longitudinal strain
HFpEF: heart failure with preserved ejection fraction
HFrEF: heart failure with reduced ejection fraction
HR: hazard ratio
LV: left ventricular
MOOSE: meta-analysis of observational studies in epidemiology
QUIPS: quality in prognosis studies
RV: right ventricular
RVEF: right ventricular ejection fraction
TAPSE: tricuspid annular plane systolic excursion

## SUPPLEMENTARY DATA

**Supplementary Table 1**. Search syntax

**Supplementary Table 2**. Echocardiographic setup of the ten eligible studies

**Supplementary Table 3**. Comparison of the reported right ventricular functional parameters

**Supplementary Figure 1**. Quality of included studies assessed using the QUIPS tool

**Supplementary Figure 2**. Funnel plot of studies assessing three-dimensional right ventricular ejection fraction as predictor of all-cause mortality and/or composite adverse cardiopulmonary events

**Supplementary Figure 3**. Funnel plot and trim-and-fill analysis of studies assessing three-dimensional right ventricular ejection fraction as predictor of all-cause mortality and/or composite adverse cardiopulmonary events

**Supplementary Figure 4**. Three-dimensional (3D) echocardiography-derived right ventricular ejection fraction (3D RVEF) and tricuspid annular plane systolic excursion (TAPSE) as predictors of all-cause mortality and/or composite adverse cardiopulmonary endpoints

**Supplementary Figure 5**. Funnel plot of studies assessing three-dimensional echocardiography-derived right ventricular ejection fraction versus tricuspid annular plane systolic excursion as predictors of all-cause mortality and/or composite adverse cardiopulmonary endpoints

**Supplementary Figure 6**. Three-dimensional (3D) echocardiography-derived right ventricular ejection fraction (3D RVEF) and fractional area change (FAC) as predictors of all-cause mortality and/or composite adverse cardiopulmonary endpoints

**Supplementary Figure 7**. Funnel plot of studies assessing three-dimensional echocardiography-derived right ventricular ejection fraction versus fractional area change as predictors of all-cause mortality and/or composite adverse cardiopulmonary endpoints

**Supplementary Figure 8**. Three-dimensional (3D) echocardiography-derived right ventricular ejection fraction (3D RVEF) and free-wall longitudinal strain (FWLS) as predictors of all-cause mortality and/or composite adverse cardiopulmonary endpoints

**Supplementary Figure 9**. Funnel plot of studies assessing three-dimensional echocardiography-derived right ventricular ejection fraction versus free-wall longitudinal strain as predictors of all-cause mortality and/or composite adverse cardiopulmonary endpoints

## DATA AVAILABILITY STATEMENT

The data underlying this article will be shared on reasonable request to the corresponding author.

