## Supplementary Tables and Figures for "Superior prognostic value of three-dimensional echocardiography-derived right ventricular ejection fraction: a meta-analysis"

**SUPPLEMENTARY MATERIAL**

**Supplementary Table 1.** Search syntax

(“3D”[All Fields] OR “3-D”[All Fields] OR “3-dimensional”[All Fields] OR “three-dimensional”[All Fields] OR “4D”[All Fields] OR “4-D”[All Fields] OR “4-dimensional”[All Fields] OR “four-dimensional”[All Fields]) AND (“Echocardiography”[MeSH Terms] OR echocardiograph*[All Fields] OR “echo”[All Fields] OR “ultrasound”[All Fields]) AND (“right ventricular”[All Fields] OR “RV”[All Fields] OR “right ventricle”[All Fields] OR “right heart”[All Fields]) AND (“ejection fraction”[All Fields] OR “EF”[All Fields] OR “RVEF”[All Fields] OR “3DRVEF”[All Fields] OR “failure”[All Fields] OR “dysfunction”[All Fields]) AND (“cardiac event”[All Fields] OR “cardiovascular event”[All Fields] OR “major cardiovascular events”[All Fields] OR “major adverse cardiovascular events”[All Fields] OR “MACE”[All Fields] OR “cardiac death”[All Fields] OR “cardiovascular death”[All Fields] OR “death”[All Fields] OR mortalit*[All Fields] OR “Mortality”[MeSH Terms] OR hazard ratio*[All Fields] OR “HR”[All Fields] OR “Cox”[All Fields] OR “Cox regression”[All Fields] OR “proportional hazards regression”[All Fields] OR “hospitalization”[All Fields] OR “Hospitalization”[MeSH Terms] OR prognos*[All Fields] OR “Prognosis”[MeSH Terms] OR surviv*[All Fields] OR “Survival Analysis”[MeSH Terms] OR predict*[All Fields])

**Supplementary Table 2. Echocardiographic setup of the ten eligible studies**

| **Study** | **Ultrasound system** | **Probe** | **3D volume rate, VPS** | **single-beat / multi-beat** | **RV-focused 3D-dataset** | **Software** | **Software version** | **Manual correction** | **FWLS software** |
| --- | --- | --- | --- | --- | --- | --- | --- | --- | --- |
| **Murata (*2016)*** | GE Vivid E9 | NA | NA | NA | NA | TomTec 4D RV-Function | NA | NA | GE EchoPAC |
| **Moceri *(2017)*** | Philips iE33 or  EPIQ 7 | X5-1 | 17.7 (16.6–18.7) | 2 beats | yes | TomTec 4D RV-Function | 2.0 | yes | NA |
| **Surkova *(2019)*** | GE Vivid E9 | 4V-D | NA | 4 to 6 beats | yes | TomTec 4D RV-Function | 2.0 | yes | NA |
| **Zhang (*2021)*** | Philips EPIQ 7C | NA | NA | single-beat | yes | 3D Auto RV, Philips | NA | yes | TomTec CPA |
| **Vîjîiac *(2021)*** | GE Vivid E9 | 4V-D | NA | 6 beats | yes | TomTec 4D RV-Function | NA | yes | GE EchoPAC |
| **Li *(2021)*** | Philips EPIQ 7C | X5-1 | NA | up to 6 beats | yes | 3D Auto RV, Philips | NA | yes | NA |
| **Meng (*2021)*** | Philips iE33 | NA | 20-35 | 4 beats | yes | TomTec 4D RV-Function | 2.0 | yes | TomTec CPA |
| **Tolvaj *(2021)*** | Philips EPIQ / GE Vivid E95 | X5-1 / 4V-D or 4Vc-D | min. 25 | Mixed single and multi-beat | yes | TomTec 4D RV-Function | 2.0 | yes | TomTec 4D RV-Function 2.0 |
| **Nabeshima *(2021)*** | Philips iE33 or EPIQ 7G/ GE Vivid7 or Vivid E95 | X3-1 or X5-1/4V-D | NA | NA | yes | TomTec 4D RV-Function | 3.0 | yes | TomTec AutoStrain RV |
| **Kitano (*2022)*** | Philips iE33 or EPIQ 7G/GE Vivid E95 | X5-1/4V-D or 4Vc-D | 23 (20–27) | Mixed single and multi-beat | yes | TomTec 4D RV-Function | 3.0 | yes | NA |

Data are presented as median (interquartile range). 3D: three-dimensional, FWLS: free wall longitudinal strain, RV: right ventricular, VPS: volumes per second

**Supplementary Table 3. Comparison of the reported right ventricular functional parameters**

| **Study** | **3D RVEF** | **TAPSE** | **RV FAC** | **RV FWLS** |
| --- | --- | --- | --- | --- |
| **Murata (*2016)*** | 43±12 | 19±4 | 31±11 | −19.7±6.4 |
| **Moceri *(2017)*** | 35.6±9.7 | 20.4±5.2 | NA | NA |
| **Surkova *(2019)*** | 48 (41-52) | 20 (16-24) | 40 (32-45) | NA |
| **Zhang *(2021)*** | 48.5±5.8 | 22.9±3.8 | 47.4±5.7 | −22.9±4.8 |
| **Vîjîiac *(2021)*** | 42±10 | 18±5 | 33±12 | −14.8±8.3 |
| **Li *(2021)*** | 37.5±9.1 | 15.5±5.2 | 28.4±8.8 | −18.6±5.5 |
| **Meng *(2021)*** | 45.6±4.7 | 20.0±3.0 | 41.5±4.5 | −20.5±3.5 |
| **Tolvaj *(2021)*** | 46.9±9 | 20.2±6.6 | 41.1±8.7 | 23.6±7 |
| **Nabeshima *(2021)*** | 48.0 (43.7-52.7) | NA | 40.1 (36.1–44.8) | 24.6 (20.7–27.8) |
| **Kitano (*2022)*** | 48 (40-54) | 16.7 (13-20.6) | NA | NA |

Data are presented as mean±SD or median (interquartile range). 3D: three-dimensional, FAC: fractional area change, FWLS: free wall longitudinal strain, RV: right ventricular, TAPSE: tricuspid annular plane systolic excursion

**Supplementary Figure 1. Quality of included studies assessed using the QUIPS tool.**

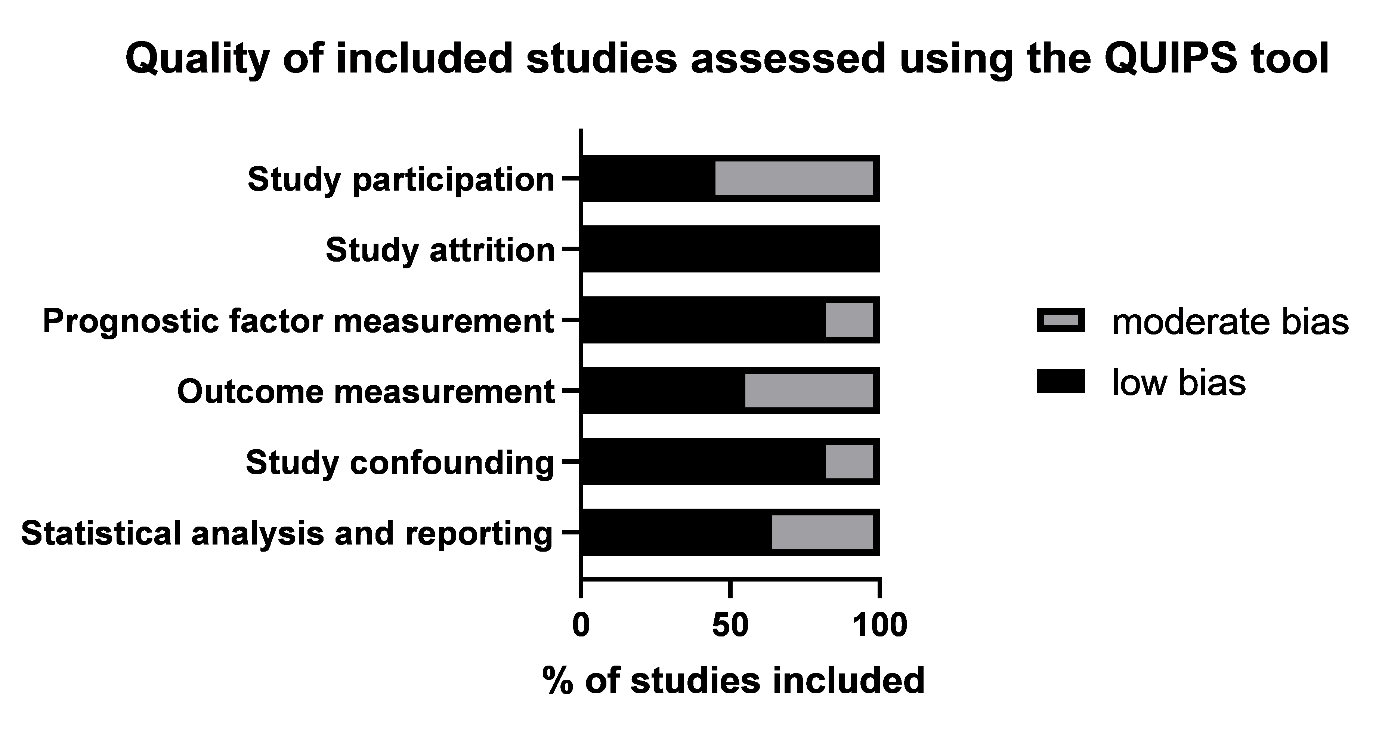

**Supplementary Figure 2. Funnel plot of studies assessing three-dimensional right ventricular ejection fraction as predictor of all-cause mortality and/or composite adverse cardiopulmonary events.**

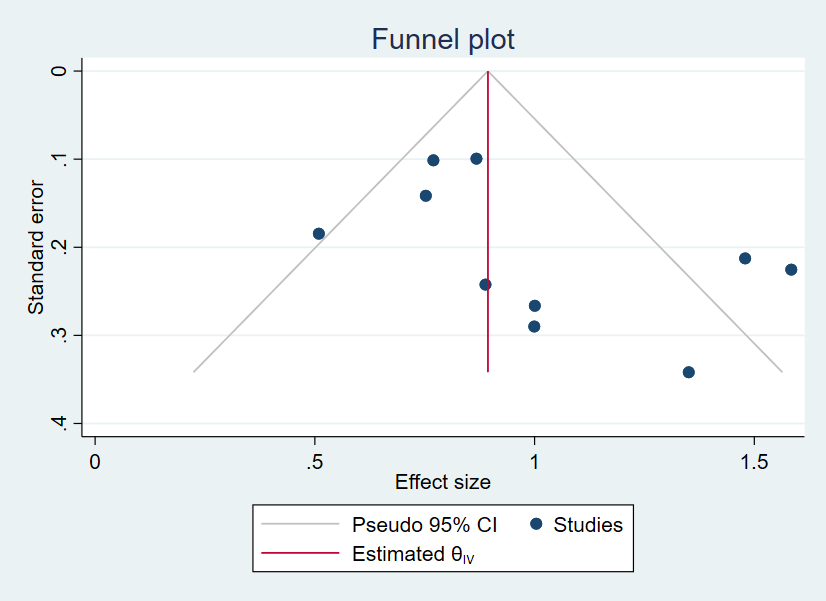

**Supplementary Figure 3. Funnel plot and trim-and-fill analysis of studies assessing three-dimensional right ventricular ejection fraction as predictor of all-cause mortality and/or composite adverse cardiopulmonary events.**

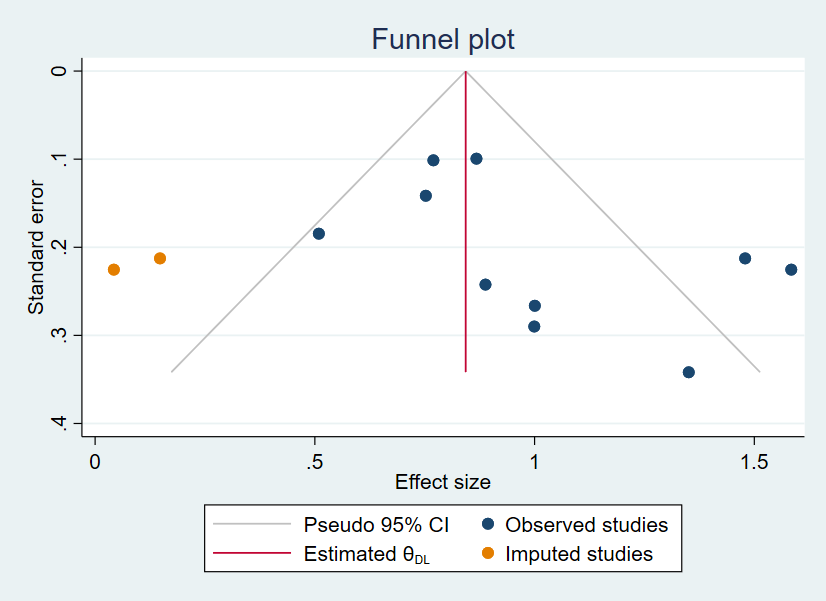

Blue dots represent the original data points (observed studies). The two orange dots represent the two imputed study estimates in accordance with Duval and Tweedie trim-and-fill analysis using the G0 estimator. Accordingly, the red line represents the bias-adjusted overall estimate.

**Supplementary Figure 4.** **Three-dimensional (3D) echocardiography-derived right ventricular ejection fraction (3D RVEF) and tricuspid annular plane systolic excursion (TAPSE) as predictors of all-cause mortality and/or composite adverse cardiopulmonary endpoints.**

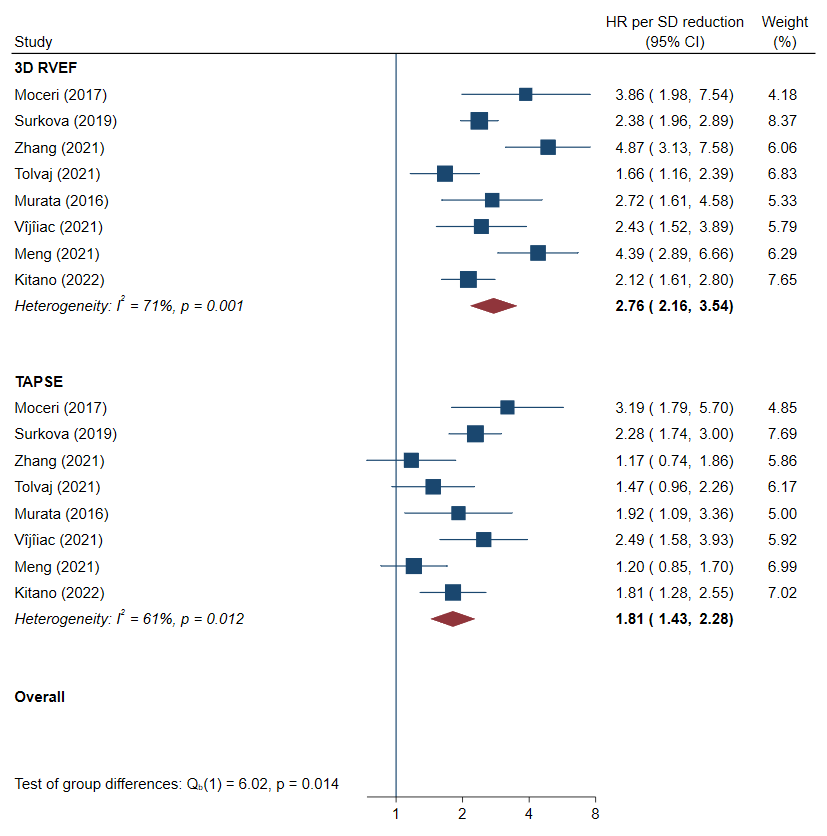

The hazard ratios (HR) are per 1 standard deviation (SD) reduction in RVEF or TAPSE according to each study. Accordingly, a HR of >1.00 means that 1 SD reduction in RVEF or TAPSE is associated with increased risk of adverse events.

CI: confidence interval

**Supplementary Figure 5.** **Funnel plot of studies assessing three-dimensional echocardiography-derived right ventricular ejection fraction versus tricuspid annular plane systolic excursion as predictors of all-cause mortality and/or composite adverse cardiopulmonary endpoints.**

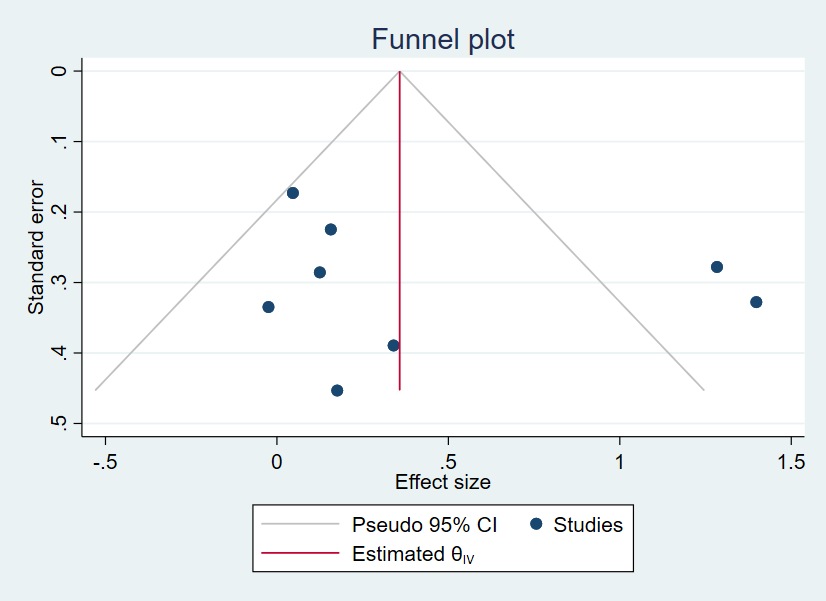

**Supplementary Figure 6.** **Three-dimensional (3D) echocardiography-derived right ventricular ejection fraction (3D RVEF) and fractional area change (FAC) as predictors of all-cause mortality and/or composite adverse cardiopulmonary endpoints.**

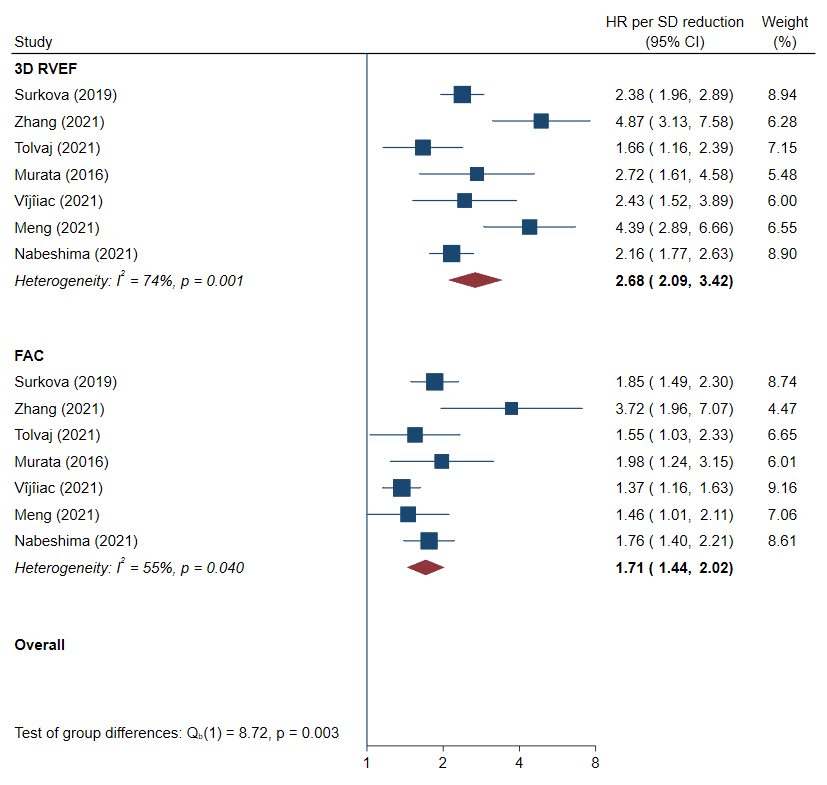

The hazard ratios (HR) are per 1 standard deviation (SD) reduction in RVEF or FAC according to each study. Accordingly, a HR of >1.00 means that 1 SD reduction in RVEF or FAC is associated with increased risk of adverse events.

CI: confidence interval

**Supplementary Figure 7. Funnel plot of studies assessing three-dimensional echocardiography-derived right ventricular ejection fraction versus fractional area change as predictors of all-cause mortality and/or composite adverse cardiopulmonary endpoints.**

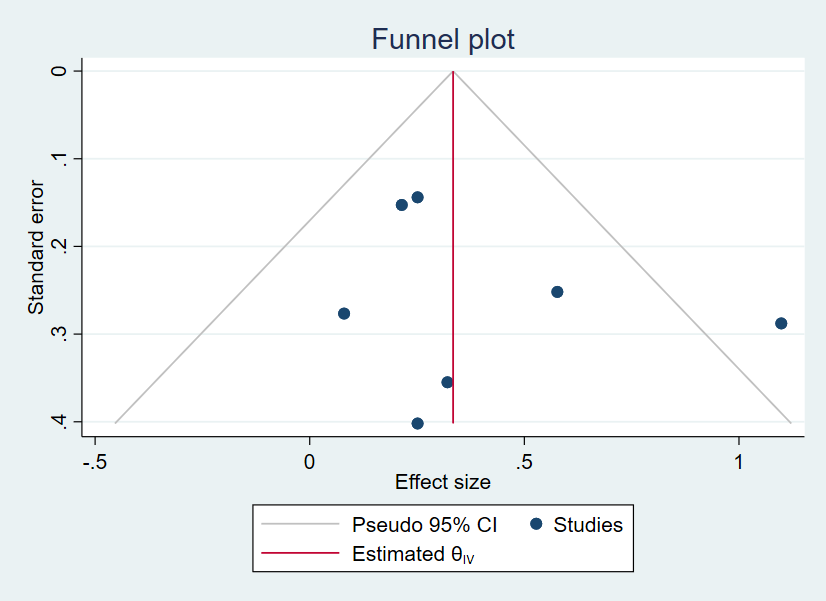

**Supplementary Figure 8. Three-dimensional (3D) echocardiography-derived right ventricular ejection fraction (3D RVEF) and free-wall longitudinal strain (FWLS) as predictors of all-cause mortality and/or composite adverse cardiopulmonary endpoints.**

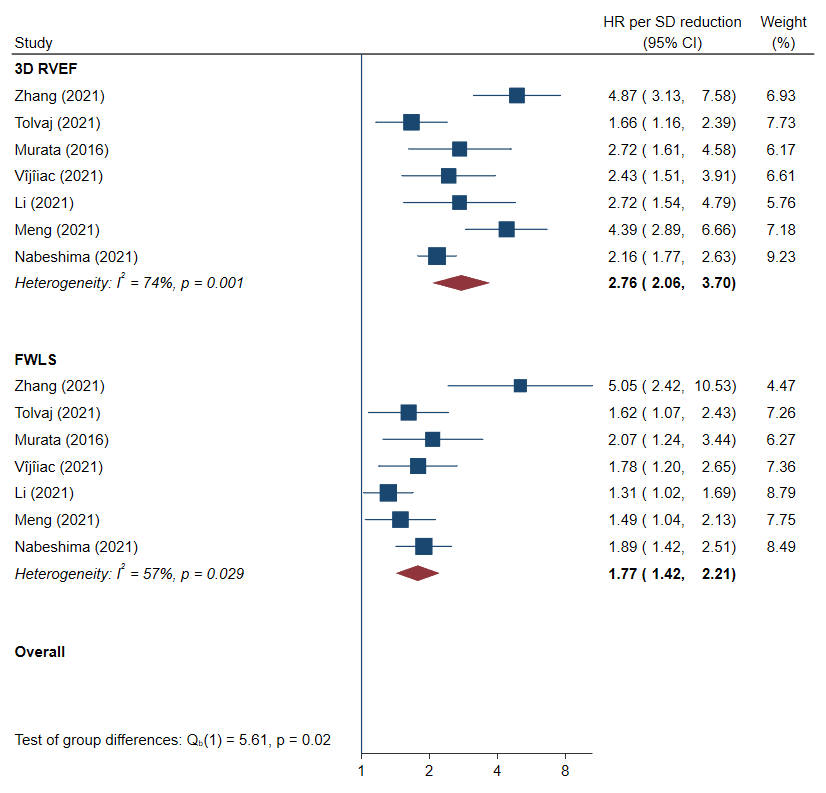

The hazard ratios (HR) are per 1 standard deviation (SD) reduction in RVEF or FWLS according to each study. Accordingly, a HR of >1.00 means that 1 SD reduction in RVEF or FWLS is associated with increased risk of adverse events.

CI: confidence interval

**Supplementary Figure 9. Funnel plot of studies assessing three-dimensional echocardiography-derived right ventricular ejection fraction versus free-wall longitudinal strain as predictors of all-cause mortality and/or composite adverse cardiopulmonary endpoints.**

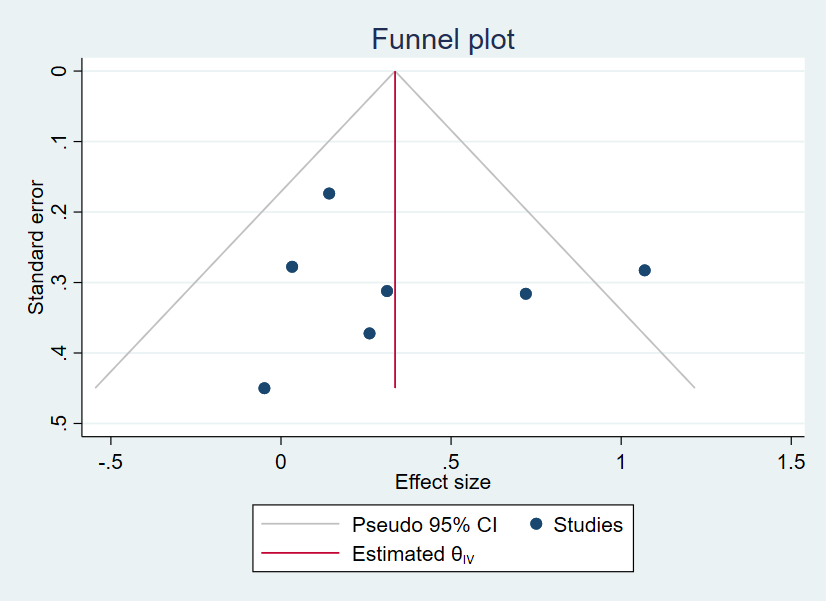
